# Agentic Artificial Intelligence for Hospital Readmission Review: A Single-Center Blinded Evaluation and Exploratory Qualitative Analysis

**DOI:** 10.64898/2026.06.17.26355917

**Authors:** Michael Gensheimer, Rishav Adhikari, Caitlin Parmer-Chow, Nancy Liu, Stephen Ma, Lisa Shieh

## Abstract

**Background:** Manual review of 30-day hospital readmissions can identify actionable quality and safety problems, but it is labor-intensive. We developed and evaluated an agentic AI workflow for evidence-grounded readmission review.

**Materials and methods:** We studied adult patients with unplanned 30-day readmission after discharge from a medicine hospitalist service at a single academic health system. An AI agent using a large language model queried a database containing notes, encounters, procedures, laboratory results, and other clinical data, and completed the same structured readmission-review rubric used by physicians. In the primary comparative evaluation, 20 randomly selected readmissions from 2025 were each reviewed by two physicians and the AI system. Blinded physician evaluators rated review quality. After rubric refinement, the AI workflow was applied to 100 recent readmissions in an exploratory expanded-cohort analysis of recurring improvement opportunities.

**Results:** In the primary comparative evaluation, the AI classified 9/20 readmissions (45%) as preventable, compared with 19/40 physician reviews (47.5%). Blinded overall quality ratings were similar for AI and physician reviews (4.35 vs. 4.20 on a 1-5 scale; mean difference 0.15, 95% CI -0.20 to 0.48; p=0.49), as were factuality/support and usefulness/actionability ratings. No AI hallucinations were identified during factuality review. Agreement on preventability and primary readmission category was low for both AI-human and human-human comparisons. The AI system cost $0.23 per chart; physician reviewers took a median of 15 minutes, corresponding to an estimated $42.43 per chart. In the exploratory expanded-cohort analysis, AI-assisted review identified recurring vulnerabilities in post-discharge follow-up plans, incomplete inpatient workups, medication-safety transitions, and indwelling-device transitions.

**Conclusions:** Agentic AI produced readmission reviews with similar blinded quality ratings to physician reviews in this small single-center primary comparative evaluation and supported identification of recurring quality-improvement themes in the exploratory expanded-cohort analysis. Preventability judgments remained variable among both AI and physicians, underscoring the need for human oversight and prospective evaluation before operational use.

**Key messages:** *What is already known on this topic:* Manual readmission review can identify modifiable quality and safety problems, but it is labor-intensive, and prior work has shown that judgments about preventability have limited interrater reliability.

*What this study adds:* In the primary comparative evaluation, an agentic AI chart-review workflow produced readmission reviews with similar quality ratings to physician reviews, and no hallucinations identified during factuality review.

*How this study might affect research, practice or policy:* AI-assisted readmission review could help health systems screen larger cohorts and identify recurring improvement opportunities, but variable preventability judgments and occasional over-attribution of care gaps demonstrate that clinician oversight remains essential.

## Introduction

Readmission soon after hospital discharge is a common problem, resulting in increased costs to the health system and impacting patient experience [1–3]. As readmission rate is considered a potential marker for quality of care, and hospitals in the United States are penalized for excess 30-day readmissions, this metric is an important focus of quality improvement programs [4]. Readmissions also matter directly to patients. In a large study, higher ratings on person-centred inpatient quality indicators, including overall hospital experience, physician communication, and medication information, were associated with lower 30-day readmission odds; readmissions also reduce home time, a patient-centered outcome tied to quality of life [5,6]. However, determining whether a readmission was preventable requires reconstructing the index hospitalization, discharge plan, and post-discharge course as well as determining causal plausibility of potential care gaps. This case-level judgment is difficult because readmissions are often multifactorial; prior work has identified common drivers of readmissions including lack of follow-up appointments after discharge, premature discharge, and patient uncertainty about who to contact after discharge [3]. A scalable review process that can identify actionable, evidence-supported contributors to readmission could therefore help health systems move from reporting readmission rates to understanding specific opportunities for improvement and targeting interventions accordingly.

Manual medical record review can provide richer clinical context than structured administrative data, but it imposes substantial burdens on health systems and can be subjective. A single-center study of externally reported inpatient quality metrics found that data collection and reporting required more than 108,000 person-hours annually [7]. Prior studies have also found substantially lower agreement for judgments about preventability than for event detection, including in adverse-event and readmission review [8–10]. A key challenge is how to make case review more scalable while preserving the clinical detail needed for fair and useful quality assessment.

The information needed for this type of review is often documented in narrative chart text rather than structured data fields, creating an opportunity for AI-assisted review. Early natural language processing systems improved detection of safety events in free-text notes beyond administrative codes, and more recent large language model (LLM) systems have shown strong agreement with manual review for clinical data abstraction and complex hospital quality measurement [11–14]. LLMs have also been used to measure guideline-concordant care and to support patient-safety event detection with traceable, expert-grounded workflows [15,16]. Most prior evaluations, however, have focused on detection, prediction, or abstraction tasks. Fewer studies have directly compared AI-generated analyses with expert human case reviews for higher-level quality-improvement tasks that require longitudinal synthesis, interpretation, and causal judgment.

Readmission review may be especially well suited to an agentic AI workflow, in which the model can search the chart, retrieve specific records, and synthesize evidence over multiple steps rather than relying on a single static chart extract. Such workflows resemble iterative clinical case review: identifying a possible issue, following leads across the chart, and integrating multiple evidence streams into a causal assessment. This approach is supported by work showing that language models can improve on multistep tasks by generating intermediate reasoning steps and interleaving reasoning with external actions or tool calls [17–19]. It may also be useful for large EHRs, where models can fail to reliably use relevant information as context length increases [20].

In this work, we developed and evaluated an agentic AI chart-review tool designed to produce evidence-grounded and actionable assessments of 30-day hospital readmissions. To our knowledge, this is among the first studies to compare AI and physician performance on case review as an interpretive quality-improvement task.

## Methods

### Patients

Data were retrieved from the Stanford STARR-OMOP database [21]. This research database contains deidentified clinical data for more than 3.4 million patients seen in the Stanford health system, including demographics, encounters, notes, procedures, labs, and follow-up information. Notes were deidentified by automatically replacing identifiers such as patient name with other randomly chosen identifiers. Dates were randomly shifted by up to 30 days, consistently across all notes and events for that patient.

Adult patients who had an unplanned readmission within 30 days of discharge in 2025-2026 were included. Only index/readmission pairs where the index hospitalization was on a general medicine service at Stanford Hospital were included. For patients with multiple readmissions within the 30-day window, only the first readmission was included. Twenty random index admissions in 2025 were selected for further analysis.

### Chart reviews

To do their chart reviews, internal medicine trained physicians and the AI chart-review system were given the patient’s chart data from the first day of the index admission to the last day of the reviewed readmission. Data included clinical notes (including radiology and pathology reports), procedures, lab results, and encounter dates/departments.

The review rubric was adapted from an existing Stanford quality-improvement survey administered to discharging physicians when a patient was readmitted within 30 days. The rubric asked reviewers to determine whether the index and readmission encounters were both true inpatient admissions, assess source-data completeness, summarize the index hospitalization, summarize the events leading to readmission, identify possible contributors to readmission, determine whether actionable care gaps were present, and judge whether the readmission was preventable.

A readmission was considered preventable only when both conditions were met: first, an actionable care gap or system failure occurred during the index hospitalization, transition of care, or post-discharge interval; and second, correcting that gap would reasonably have prevented or delayed the readmission given the patient’s clinical trajectory, engagement with care, and social circumstances. Care gaps could be documented without classifying the readmission as preventable if the gap was not causal. The primary reason for readmission was categorized into one of the following categories, based on the findings of Auerbach et al. and our operationally-used readmission survey: discharge planning, care coordination/follow-up; diagnostic, medication, or monitoring issues; patient education, social determinants of health/advance care planning; unavoidable readmission; or other [3].

For the physician reviewers, a chart viewer web application was created that allowed them to browse and search the chart data (Supplementary Figure S1).

For each patient chart, we had two physician reviewers and the AI system complete the review rubric.

### AI chart-review system

For automated chart abstraction, we implemented an AI agent that was able to browse and search the patient’s chart to extract and analyze relevant information. We used the open source OpenAI Agents software development kit with the Gemini 3.1 Pro Preview LLM (2/19/2026 version). The LLM was run under Stanford’s business associate agreement with Google, which allows use of protected health information and prohibits Google from training using Stanford data.

For each patient, the agent received the index and readmission encounter identifiers and queried a read-only SQLite database export of the relevant time period of the patient’s deidentified STARR-OMOP chart. The database included encounters, notes, procedures, laboratory results, and other clinical results. The agent could only execute read-only SQL SELECT queries.

The agent prompt requested a structured output following the same review rubric used by human reviewers. Before free-form chart exploration, the agent was required to retrieve key source documents, including the index admission H&P, discharge summary, discharge instructions, and readmission documents (ED provider note, inpatient H&P, discharge documentation) when available. It was then instructed to perform focused chart review of the index admission and post-discharge interval. The prompt also included information about existing Stanford quality initiatives (see Supplement for full prompt). Three AI reviews were performed per chart and then synthesized into a single final output using a separate LLM combiner that prioritized evidence-supported claims. This step was performed because in early testing, when there was a key fact located in only one place in the chart, the AI agent usually found it but not always. Repeating the AI abstraction three times reduces the risk of missing a key fact.

A physician iteratively refined the AI instructions using patients not included in the primary comparative evaluation. Refinement focused on reducing unsupported speculation, enforcing explicit evidence citation, clarifying the distinction between care gaps and preventability, and aligning the AI rubric with the physician-review workflow. After this development phase, three physician reviewers and the AI system reviewed one calibration patient; the physicians then met to harmonize grading conventions and further refine the AI instructions before applying the final rubric to the primary comparative evaluation sample (see Supplemental Methods).

## Statistical Analysis

We summarized patient characteristics and review outputs using descriptive statistics. For structured review items, including readmission preventability and primary reason for readmission, we calculated percent agreement and Cohen’s kappa for AI-human pairs and for human-human pairs.

We had physician raters score the quality of each response, blinded to the identities of the reviewers. The raters had access to the EHR data but were not expected to look through the whole chart; instead they were asked to focus on evaluating the factuality of the analysis statements. They completed Likert scale scores for overall quality, factuality/support, usefulness/actionability, and reviewer identity (physician vs. AI) and were also able to give free-text comments. To compare quality scores between AI and human reviews, we calculated patient-level paired differences by subtracting the mean score of the physician reviewers from the corresponding AI review score. Statistical significance was assessed using a two-sided exact paired sign-flip permutation test, with 95% confidence intervals for the mean difference constructed using bootstrap percentile estimation.

We also calculated similarity between different reviews for the same patient. For free-text items, including summaries of the index hospitalization, events leading to readmission, reasons for readmission, and preventability explanation, pairwise similarity was rated on a 5-point Likert scale. Similarity ratings were performed by an LLM using a prespecified rubric. To validate this approach, a physician independently scored a subset of text pairs, and agreement between physician and LLM ratings was assessed using quadratic weighted kappa and within-1-point agreement.

For each patient, AI-human similarity was defined as the mean of the two AI-physician similarity scores, and human-human similarity was defined as the similarity score between the two physician reviewers. Patient-level AI-human and human-human scores were compared using two-sided exact paired sign-flip permutation tests. The same patient-level approach was used to compare AI and physician word counts.

After the primary comparative evaluation, we refined the rubric and AI prompt and applied the revised AI workflow to the 100 most recent eligible readmissions from the same clinical services for an exploratory expanded-cohort analysis. In contrast to the primary comparative evaluation, to mimic real-world usage, a patient could be included more than once in the 100-readmission cohort if they had multiple readmissions during the target timeframe. This exploratory expanded-cohort analysis was used to assess recurring care gaps and improvement opportunities. We conducted a qualitative content analysis of the 100 reviews. A mixed deductive-inductive codebook was developed, using readmission driver categories, guideline adherence items, prevention opportunities, and patient factors as initial deductive codes. To calibrate the codebook and ensure conceptual alignment, two physicians independently coded an initial subset of 10 cases. The reviewers then met to compare their findings, resolve discrepancies through consensus, and refine the code definitions. The primary reviewer evaluated the remaining 90 cases using this finalized framework. An LLM (Gemini Flash 3.5) was used to draft preliminary codes, which were reviewed and edited by the reviewer.

## Results

The primary comparative evaluation was performed on 20 randomly selected patients who had a readmission within 30 days of an index hospitalization on a medicine hospitalist service in 2025. Table 1 shows patient characteristics.

**Table 1.**
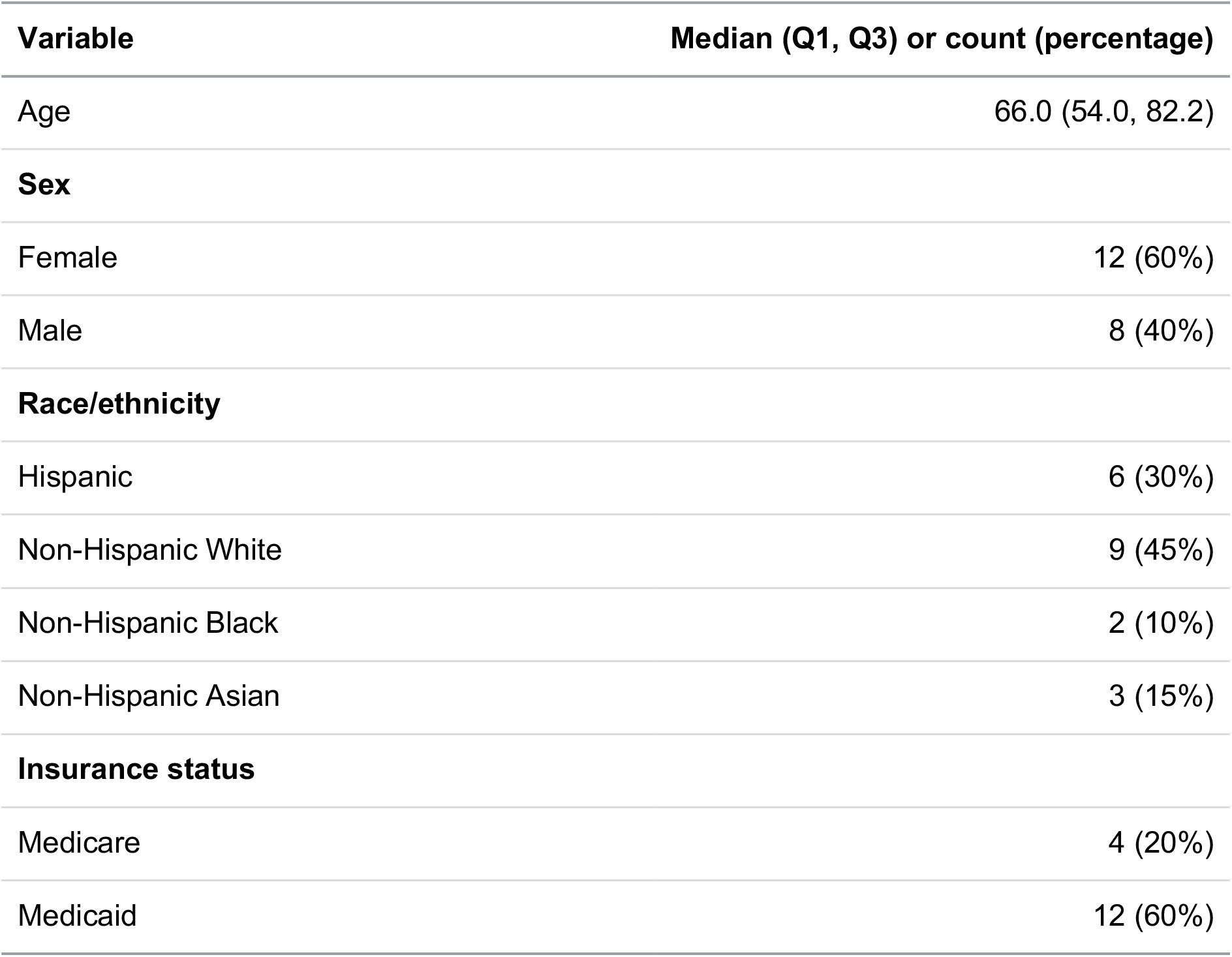
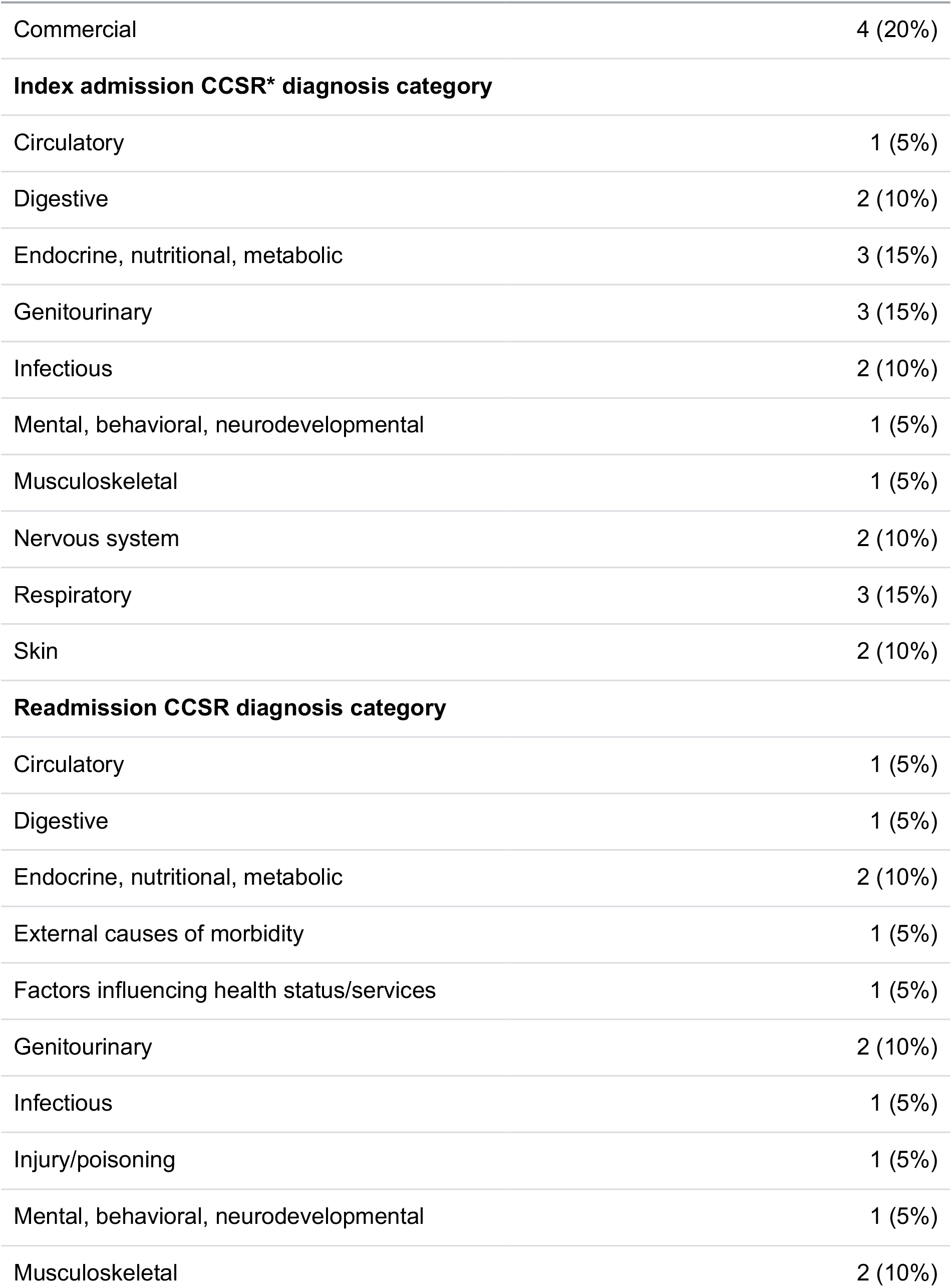
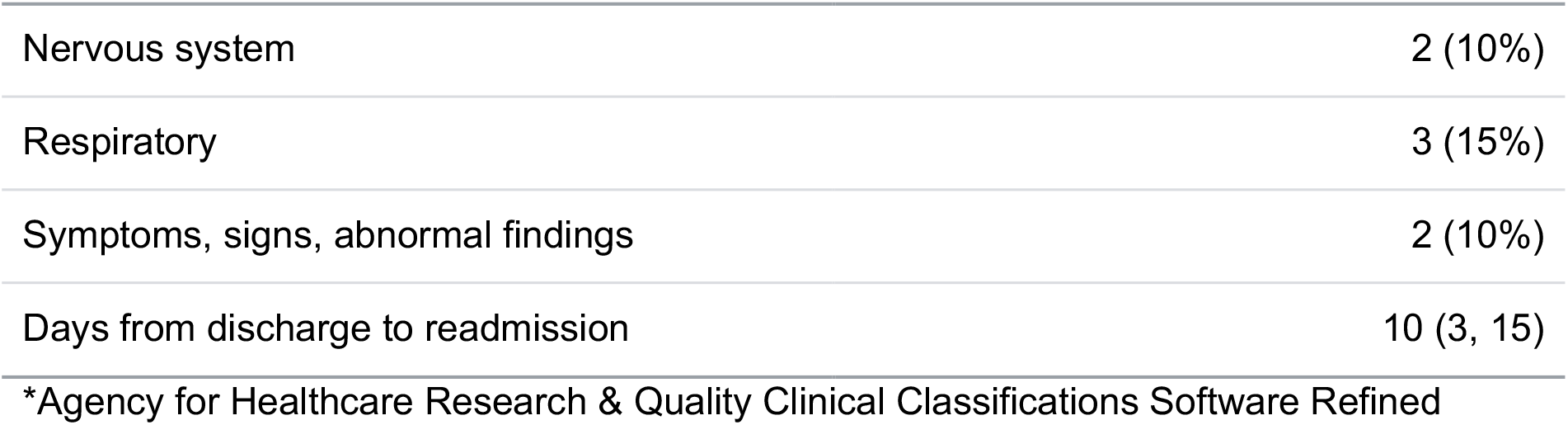
Patient characteristics (n=20)

| Variable | Median (Q1, Q3) or count (percentage) |
| --- | --- |
| Age | 66.0 (54.0, 82.2) |
| **Sex** |  |
| Female | 12 (60%) |
| Male | 8 (40%) |
| **Race/ethnicity** |  |
| Hispanic | 6 (30%) |
| Non-Hispanic White | 9 (45%) |
| Non-Hispanic Black | 2 (10%) |
| Non-Hispanic Asian | 3 (15%) |
| **Insurance status** |  |
| Medicare | 4 (20%) |
| Medicaid | 12 (60%) |

| **Variable** | **Median (Q1, Q3) or count (percentage)** |
| --- | --- |
| Commercial | 4 (20%) |
| **Index admission CCSR\* diagnosis category** |  |
| Circulatory | 1 (5%) |
| Digestive | 2 (10%) |
| Endocrine, nutritional, metabolic | 3 (15%) |
| Genitourinary | 3 (15%) |
| Infectious | 2 (10%) |
| Mental, behavioral, neurodevelopmental | 1 (5%) |
| Musculoskeletal | 1 (5%) |
| Nervous system | 2 (10%) |
| Respiratory | 3 (15%) |
| Skin | 2 (10%) |
| **Readmission CCSR diagnosis category** |  |
| Circulatory | 1 (5%) |
| Digestive | 1 (5%) |
| Endocrine, nutritional, metabolic | 2 (10%) |
| External causes of morbidity | 1 (5%) |
| Factors influencing health status/services | 1 (5%) |
| Genitourinary | 2 (10%) |
| Infectious | 1 (5%) |
| Injury/poisoning | 1 (5%) |
| Mental, behavioral, neurodevelopmental | 1 (5%) |
| Musculoskeletal | 2 (10%) |
| Nervous system | 2 (10%) |
| Respiratory | 3 (15%) |
| Symptoms, signs, abnormal findings | 2 (10%) |
| Days from discharge to readmission | 10 (3, 15) |

\*Agency for Healthcare Research & Quality Clinical Classifications Software Refined

Two physicians and the AI system reviewed each chart. Patterns of responses are shown in Supplemental Table S1. The AI rated 45% of readmissions (9/20) as preventable; on average, humans rated 47.5% of readmissions as preventable (19/40). AI answers for the four major free-text fields (index hospitalization summary, readmission lead-up, readmission reasons, and preventability explanation) were significantly longer than human answers (99.3 vs. 48.6 mean words overall, p<0.001, Supplemental Table S2).

### Review quality

For each patient, an attending hospitalist rated the quality of the three reviews (two human, one AI) on a 1-5 Likert Scale. The rater was blinded to the identity of the reviewer, including whether the review was completed by AI or a physician. Results are shown in Table 2. Overall quality score was similar for AI and humans: 4.35 vs. 4.20, mean difference 0.15 (-0.20, 0.48) slightly favoring the AI, p=0.49).

**Table 2.** Physicians’ quality ratings of case reviews for 20 patients (20 reviews done by AI, 40 by physicians).

| Item | Mean AI score (1-5) | Mean Human score (1-5) | Mean AI - human score (95% CI) | p value |
| --- | --- | --- | --- | --- |
| Overall quality | 4.35 | 4.20 | 0.15 (-0.20, 0.48) | 0.49 |
| Factuality/<br>support | 4.40 | 4.35 | 0.05 (-0.35, 0.45) | 0.91 |
| Usefulness/<br>actionability | 4.25 | 4.00 | 0.25 (-0.25, 0.725) | 0.39 |

No AI hallucinations were identified during physician review. Evaluators also recorded their guess as to the identity of each case reviewer (AI vs. human). They were correct in 60/60 cases (100%).

We identified themes from qualitative analysis of the case reviews. The AI system’s answers were longer and more structured; for instance its discussion of reasons for readmission frequently used bulleted lists broken down by reason category. Some human responses were comprehensive but some were very short, leaving out important details or citations that would support the argument. The AI system was more likely to cite specific quotes from notes or other chart data. The AI system was also more likely to recommend system-level changes such as increased use of a meds-to-beds program or mandating stricter outpatient visit scheduling guidelines. While physician reviewers relied most heavily on data from discharge summaries and re-admission history and physical notes, the AI was more likely to identify care gaps that were visible only in other clinical documents. Finally, it sometimes suggested that a care gap occurred such as missed outpatient visit scheduling, when the human reviewers noted that this was mainly because of the patient refusing care due to substance use disorder or other issues. Highlights of results from three selected patients are in Supplemental Table S3.

We also examined the similarity of key structured and unstructured fields from AI versus human chart reviews; results are in the Supplement.

### Cost

The cost per chart to run the AI system was 23 cents. The human case reviewers took a median of 15 minutes to review each chart (IQR 11-15, 95% CI 12.7, 15.8). Based on a median total compensation for hospitalists of $353,002 and a typical work week of 40 hours, this indicates an estimated human review cost of $42.43 per chart [22], for a human:AI cost ratio of 185.

### Exploratory Expanded-Cohort Analysis

Based on the results of the primary comparative evaluation, we made refinements to the rubric and AI chart-review prompt. The AI system was now allowed to give multiple categories of care gaps, but it was still required to list the primary reason for readmission. The AI system was also able to list actionable improvement opportunities and the owner for each (hospitalist service, ED, etc.). Rather than a binary determination of readmission preventability, the AI system was instructed to judge preventability using a 5 point scale. Finally, it was instructed to use structured citations to the chart to make it easier to verify its claims.

For the exploratory expanded-cohort analysis, we then ran this updated AI system on the most recent 100 readmissions from the same services studied in the primary comparative evaluation; characteristics are shown in Supplemental Table S6. Informal physician review of a subset of the AI responses found similar performance as in the primary comparative evaluation, but formal performance scoring was not done for the expanded-cohort analysis. The AI classified 60/100 readmissions as definitely, probably, or possibly preventable. Care gap categories, actionability levels, and improvement stakeholders are summarized in Supplemental Tables S7-S9.

A physician performed qualitative analysis of the 100 AI reviews by assigning codes and summarizing themes. Codes were refined through a second physician review of the first 10 cases. A total of 515 codes were assigned; the most common are shown in Supplemental Table S10. Themes are shown in Table 3. Four primary systemic vulnerabilities were identified: post-discharge monitoring and handoff failures, incomplete inpatient workups, medication safety transition errors, and indwelling device issues.

**Table 3.** Themes from qualitative analysis of 100 readmission cases in the exploratory expanded-cohort analysis.

| Theme | Patient examples | Potential system interventions |
| --- | --- | --- |
| **Post-discharge surveillance and handoff failures**<br>High-risk patients decompensated when follow-up systems did not match their clinical needs. Gaps included virtual visits when examination or labs were needed, missed surveillance calls because of EHR classification logic, lack of ownership for abnormal post-discharge labs or vital signs, incomplete handoff to dialysis or facility clinicians, and care plans that relied on unavailable phone access or unsupported home health arrangements. | • A patient with hepatic hydrothorax had an in-person follow-up converted to telehealth, delaying volume assessment before recurrent hypoxemia<br>• A patient discharged on intensified heart failure therapy had early severe AKI and hypotension seen across several outpatient contacts without medication adjustment | • Require explicit ownership for post-discharge labs, weights, and vital signs<br>• Fix outreach exclusions for high-risk groups and assisted living dispositions<br>• Include structured handoff fields for dialysis centers, facilities, home health agencies, and patients with communication barriers |
| **Incomplete treatment or deferred diagnostic and procedural workup**<br>Some readmissions followed discharge before resolution of active high-risk conditions or before definitive diagnostic or procedural plans were complete. Patients were left in a vulnerable interval after incomplete inpatient evaluation, deferred procedures, or delayed specialty triage. | • A patient with malignant pleural effusion had rapid pre-discharge reaccumulation but no immediate drainage or scheduled pulmonology appointment, returning in respiratory distress<br>• A patient with severe gastrointestinal bleeding had a limited colonoscopy because of poor preparation and was discharged before repeat evaluation, returning with recurrent severe anemia | • Establish discharge criteria for active high-risk conditions requiring either completed inpatient management or a confirmed rapid outpatient procedure, test, or specialty appointment |
| **Medication safety transition failures**<br>Medication-related readmissions arose from discharge reconciliation errors, high-risk prescribing, inadequate medication access planning, confusing instructions, and missed or ineffective transitions-of-care pharmacist review. High-risk areas included insulin, opioids, diuretics, and bowel regimens. | • A patient with end-stage renal disease was discharged on an aggressive insulin sliding scale and returned with severe hypoglycemia<br>• A patient with recent venous thromboembolism had anticoagulation held even after minor bleeding resolved and returned with ischemic stroke | • Mandate transitions-of-care pharmacist review for patients discharged on high-risk medications or complex medication changes<br>• Use structured discharge medication checks for anticoagulation restart plans, insulin safety, opioid and bowel regimens, diuretic hold parameters |
| **Indwelling device transition** | • A patient with biliary stents developed bacteremia and | • Use standardized device-specific discharge instructions |
| **vulnerabilities**<br>Patients discharged with biliary drains, feeding tubes, nephrostomy tubes, or other devices were vulnerable when outpatient procedures, device-care instructions, medication formulations, or warning-sign guidance were not tightly coordinated. | hepatic abscess after outpatient liver biopsy without antibiotic prophylaxis<br>• A patient discharged to a facility with a new feeding tube was prescribed numerous crushed solid medications, contributing to tube clogging and rupture | with teach-back, warning signs, and direct specialty contact pathways |

As a result of this analysis, the hospitalist service is considering several potential process improvements including modifying insulin sliding scale templates, improving reliability of glucagon prescriptions, better quality assurance of discharge instructions, and improving outpatient visit scheduling prior to discharge.

## Discussion

We developed and evaluated an agentic AI chart-review system for hospital readmissions. This task is challenging and requires more than extraction of facts from the medical record. The system had to reconstruct the index hospitalization, follow the post-discharge course, identify plausible contributors to readmission, and judge whether any care gaps were actionable and causally related to the outcome. In the primary comparative evaluation, AI reviews had similar overall quality, factual support, and usefulness scores compared with physician reviews, and no hallucinations were identified. Mean AI-physician free-text similarity scores were comparable overall to the physician-physician values, and were significantly higher for the index hospitalization summary and readmission lead-up summary fields. At the same time, agreement on preventability and primary readmission category was low for both AI-human and human-human comparisons, suggesting that case review requires weighing uncertain and competing explanations. In the exploratory expanded-cohort analysis, the AI workflow produced a set of recurring, actionable themes that are now informing quality-improvement work.

### Prior work and interpretation

These findings add to a growing literature showing that language models can support clinical documentation review and quality measurement. Prior studies have shown strong performance for LLM-based abstraction of structured data elements from the EHR and for quality-measure reporting [13,14], while earlier natural language processing systems improved surveillance for patient-safety events in narrative notes [11,12]. Other work has evaluated LLMs for guideline-concordance assessment, patient-safety event detection, and clinical documentation summarization [15,16,23]. Our study differs by directly comparing AI output with physician case reviews for readmission analysis, where the task is not only to describe events but also to make a causal assessment about whether different care might reasonably have prevented the readmission. This is closer to the work of a morbidity and mortality conference or quality review committee than to a conventional abstraction or prediction task, and it tests whether AI can contribute to the interpretive layer of quality improvement that usually depends on expert human reviewers.

The agentic design was important for this use case. Readmission analysis often depends on details that are not contained in a single discharge summary or readmission H&P, such as outpatient messages, follow-up visits, and post-discharge laboratory results. A workflow that allowed the model to search the chart, retrieve source documents, and pursue leads across the index admission and gap period more closely matched how clinicians perform case review. This design is consistent with broader evidence that LLMs can perform better on multistep tasks when they generate intermediate reasoning steps and use external tools [17–19], and with emerging work on AI agents for EHR reasoning and data retrieval [24–27]. This strategy also helps address a practical limitation of long-context EHR review: relevant information can be buried in a large record and may not be reliably used if simply placed in a long prompt [20].

The AI reviews were easily distinguishable from physician reviews. Blinded evaluators correctly identified the reviewer type in every case, potentially because AI responses were longer, more structured, and more consistently cited supporting evidence. The structured and source-cited style may be valuable for quality-improvement workflows, where reviewers need to audit claims and compare patterns across cases. On the more negative side, AI reviews sometimes framed a process deviation as a care gap even when human reviewers judged that the patient was unlikely to have accepted or benefited from the intervention because of severe illness, substance use, limited engagement with care, or other contextual factors. It is possible that LLMs are trained on more idealized material such as published educational cases rather than real-world EHR data, and that this contributes to difficulty recognizing patient factors as substantial drivers of poor outcomes and readmissions.

The low agreement on preventability that we observed was consistent with prior studies of adverse-event and readmission review showing substantially worse agreement for preventability judgments than for event detection [8–10]. Readmissions are often multifactorial, and counterfactual judgments are intrinsically uncertain: a patient may have had suboptimal follow-up, medication confusion, progressive disease, and limited social support, but reviewers may reasonably disagree about whether any single improvement would have prevented hospitalization. Auerbach and colleagues found that 26.9% of readmissions were potentially preventable after case review, and identified contributors such as inadequate symptom treatment, insufficient medication monitoring, delayed follow-up, unclear instructions about whom to contact, need for additional home services, and premature discharge [3]. The themes identified in our exploratory expanded-cohort analysis were similar, especially post-discharge monitoring failures, incomplete inpatient workups, medication-safety transition errors, and device-related vulnerabilities.

### Implications

The exploratory expanded-cohort analysis illustrates the operational potential of the AI-assisted approach. At low cost, the AI system generated case-level analyses that could be aggregated into recurring themes, including discharge-instruction quality and follow-up scheduling. While this review should not replace clinical judgment or formal event investigation, it could help health systems screen larger cohorts, identify recurring vulnerabilities, and prioritize cases for human review.

This study has several limitations. It was performed at a single academic health system, focused on hospitalist readmissions, and included only 20 patients in the primary comparative evaluation. The exploratory expanded-cohort analysis was not formally evaluated against physician reviews. Because this analysis relied on AI classifications that may over-attribute the impact of care gaps, these findings should be interpreted as hypothesis-generating themes rather than estimates of the true prevalence of preventable readmissions or care gaps. The study also did not compare the agentic workflow with simpler LLM approaches.

Future work should evaluate AI readmission review prospectively and test whether AI-generated findings lead to measurable improvements in patient outcomes. AI review tools may be useful not only after readmission but before discharge, by checking whether discharge summaries, instructions, medication plans, and follow-up arrangements are consistent and aligned with guidelines. If developed with appropriate safeguards, agentic chart review could help health systems move from retrospective measurement toward continuous learning about the processes that place patients at risk.

## Data Availability

Detailed patient-level data contain sensitive protected health information and cannot be shared. Aggregate data will be shared on reasonable request.

## Ethics Approval

The project was submitted to the Stanford IRB and determined to be quality improvement rather than research.

## Patient and Public Involvement

Patients and members of the public were not involved in the design, conduct, reporting, or dissemination planning for this study.

## Funding

This study received no specific funding.

## Competing Interests

The authors declare no competing interests.

## Notes

### Competing Interest Statement

The authors have declared no competing interest.

## References

1 Jencks SF, Williams MV, Coleman EA. Rehospitalizations among patients in the medicare fee-for-service program. New England Journal of Medicine. 2009;360:1418–28. doi: 10.1056/NEJMsa0803563

2 Walraven C van, Bennett C, Jennings A, et al. Proportion of hospital readmissions deemed avoidable: A systematic review. Canadian Medical Association Journal. 2011;183:E391–402. doi: 10.1503/cmaj.101860

3 Auerbach AD, Kripalani S, Vasilevskis EE, et al. Preventability and causes of readmissions in a national cohort of general medicine patients. JAMA Internal Medicine. 2016;176:484–93. doi: 10.1001/jamainternmed.2015.7863

4 Lachar J, Avila CJ, Qayyum R. The long-term effect of financial penalties on 30-day hospital readmission rates. The Joint Commission Journal on Quality and Patient Safety. 2023;49:521–8. doi: 10.1016/j.jcjq.2023.06.001

5 Kemp K, Steele B, Ahmed S, et al. Person-centred quality indicators are associated with unplanned care use following hospital discharge. BMJ Open Quality. 2024;13:e002501. doi: 10.1136/bmjoq-2023-002501

6 Tang AB, Solomon N, Chiswell K, et al. Home-time, mortality, and readmissions among patients hospitalized with heart failure: A baseline prior to IMPLEMENT-HF. Circulation: Heart Failure. 2024;17. doi: 10.1161/CIRCHEARTFAILURE.124.011795

7 Saraswathula A, Merck SJ, Bai G, et al. The volume and cost of quality metric reporting. JAMA. 2023;329:1840–7. doi: 10.1001/jama.2023.7271

8 Klein DO, Rennenberg RJMW, Koopmans RP, et al. Adverse event detection by medical record review is reproducible, but the assessment of their preventability is not. PLOS ONE. 2018;13:e0208087. doi: 10.1371/journal.pone.0208087

9 Trickey AW, Wright JM, Donovan J, et al. Interrater reliability of hospital readmission evaluations for surgical patients. American Journal of Medical Quality. 2017;32:201–7. doi: 10.1177/1062860615623854

10 Galen LS van, Cooksley T, Merten H, et al. Physician consensus on preventability and predictability of readmissions based on standard case scenarios. The Netherlands Journal of Medicine. 2016;74:434–42.

11 Murff HJ, FitzHenry F, Matheny ME, et al. Automated identification of postoperative complications within an electronic medical record using natural language processing. JAMA. 2011;306:848–55. doi: 10.1001/jama.2011.1204

12 Ozonoff A, Milliren CE, Fournier K, et al. Electronic surveillance of patient safety events using natural language processing. Health Informatics Journal. 2022;28:14604582221132429. doi: 10.1177/14604582221132429

13 Ge J, Li M, Delk MB, et al. A comparison of a large language model vs manual chart review for the extraction of data elements from the electronic health record. Gastroenterology. 2024;166:707–709.e3. doi: 10.1053/j.gastro.2023.12.019

14 Boussina A, Krishnamoorthy R, Quintero K, et al. Large language models for more efficient reporting of hospital quality measures. NEJM AI. 2024;1. doi: 10.1056/aics2400420

15 Bannett Y, Gunturkun F, Pillai M, et al. Applying large language models to assess quality of care: Monitoring ADHD medication side effects. Pediatrics. 2024;155. doi: 10.1542/peds.2024-067223

16 Trujillo D, Wang D, Bahr N, et al. A medically grounded LLM agent-based tool to detect patient safety events in medical records. medRxiv. Published Online First: 2025. doi: 10.64898/2025.12.16.25342438

17 Wei J, Wang X, Schuurmans D, et al. Chain-of-thought prompting elicits reasoning in large language models. 2022.

18 Yao S, Zhao J, Yu D, et al. ReAct: Synergizing reasoning and acting in language models. 2022.

19 Schick T, Dwivedi-Yu J, Dessì R, et al. Toolformer: Language models can teach themselves to use tools. 2023.

20 Liu NF, Lin K, Hewitt J, et al. Lost in the middle: How language models use long contexts. 2023.

21 Callahan A, Ashley E, Datta S, et al. The stanford medicine data science ecosystem for clinical and translational research. JAMIA Open. 2023;6:ooad054. doi: 10.1093/jamiaopen/ooad054

22 Frost M. Earning what you’re worth. ACP Hospitalist. Published Online First: 2023.

23 Grolleau F, Liang AS, Keyes T, et al. Physician-reported safety outcomes of AI-generated hospital course summaries. JAMA Network Open. 2026;9:e2616556. doi: 10.1001/jamanetworkopen.2026.16556

24 Shi W, Xu R, Zhuang Y, et al. EHRAgent: Code empowers large language models for few-shot complex tabular reasoning on electronic health records. Proceedings of the 2024 conference on empirical methods in natural language processing. Association for Computational Linguistics 2024:22315–39.

25 Chen X, Jin Y, Mao X, et al. RareAgents: Autonomous multi-disciplinary team for rare disease diagnosis and treatment. Proceedings of the AAAI Conference on Artificial Intelligence. 2026;40:101–9. doi: 10.1609/aaai.v40i1.36969

26 Lee K, Hong S, Park J, et al. EMR-AGENT: Automating cohort and feature extraction from EMR databases. 2025.

27 Xu G, Li X, Chen Y, et al. A comprehensive survey of AI agents in healthcare. Journal of Biomedical Informatics. 2026;179:105045. doi: 10.1016/j.jbi.2026.105045

