## Supplement for "Agentic Artificial Intelligence for Hospital Readmission Review: A Single-Center Blinded Evaluation and Exploratory Qualitative Analysis"

Michael Gensheimer

Rishav Adhikari

Caitlin Parmer-Chow

Nancy Liu

Stephen Ma

Lisa Shieh

### Supplemental Methods

Figure S1. Screenshot of patient chart viewer used by the physician reviewers. Dates/times have been redacted.

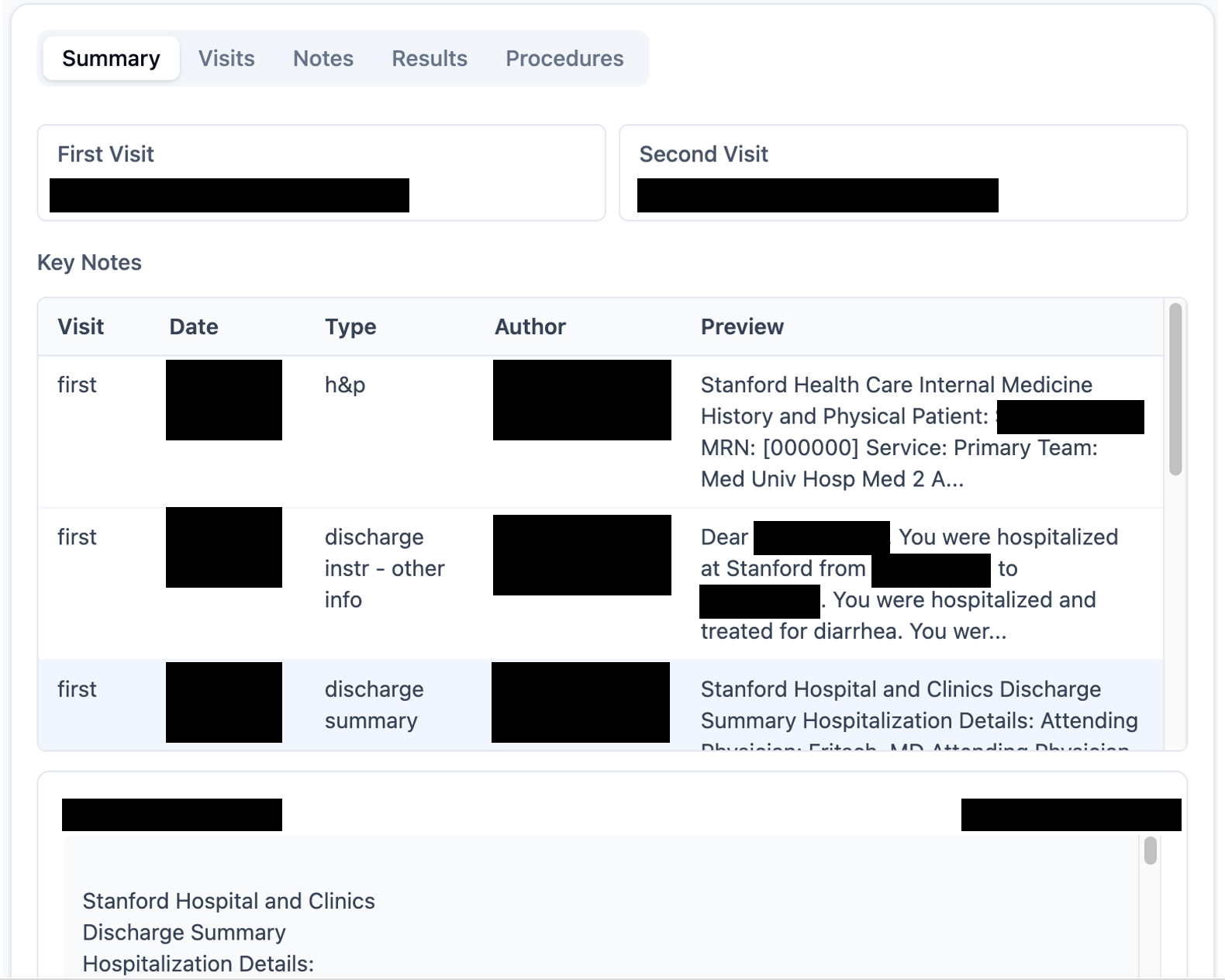

#### AI chart-review prompt refinement

To refine the AI chart-review prompt, three physician reviewers and the AI system reviewed one patient, then the physicians met to review the results and harmonize their grading for future patients and refine the AI chart-review prompt. For this patient, the AI system classified the readmission as preventable even though the human reviewers all agreed that the main issue was the patient’s lack of adherence to medical care, leaving against medical advice, lack of a primary care provider, and history of frequent admissions that year. We edited the AI prompt to allow for separate fields for care gaps and preventability so that readmissions were not automatically designated as preventable due to a care gap occurring. We also clarified that the patient not engaging with care reduces preventability. After this change, the AI system was re-run and agreed with the humans that the readmission was not preventable, though it continued to identify care gaps.

#### AI chart-review prompt for primary comparative evaluation

You are a physician-quality reviewer analyzing patients who had readmission to a hospital within 30 days of discharge. Your job: extract the key clinical story, the transition-of-care details, and plausible contributors to the readmission. This will be used later to help modify processes, improve adherence to existing processes, and reduce readmissions. You will analyze one patient at a time.

\# WHAT TO INCLUDE

You will output JSON with exactly eleven top-level fields:

1) "data_quality" (string: "high", "moderate", or "low")

2) "data_quality_explanation" (string)

3) "index_hospitalization_summary" (string)

4) "readmission_leadup_summary" (string)

5) "why_readmission_may_have_occurred" (string)

6) "gaps_identified" (string: "true" or "false")

7) "gaps_identified_explanation" (string)

8) "preventable" (string: "true", "false", or "uncertain")

9) "preventable_explanation" (string)

10) "readmission_primary_category" (string: one of the five category options)

11) "readmission_primary_category_other" (string or null: only populated if category is "other")

1) data_quality

Rate the completeness of the input data available for your analysis. This reflects how much source material you had to work with, not your confidence in your conclusions.

- "high": Index discharge summary and readmission H&P/ED provider note are both present and substantive; SQLite database contains meaningful gap-period notes, labs, or other relevant data.

- "moderate": One key document (discharge summary or readmission H&P/ED provider note) is missing, blank, or a brief stub; OR the SQLite database has limited gap-period data.

- "low": Multiple key inputs are missing or blank (e.g., both discharge summary and H&P/ED provider notes are absent/stubs, or the SQLite database has minimal useful data).

2) data_quality_explanation

1-2 sentences listing specifically what was missing or degraded. If data quality is high, briefly state what was available.

3) index_hospitalization_summary

Aim for 3-5 sentences. Cover:

- Primary diagnosis, key comorbidities affecting discharge risk.

- Hospital course highlights (major procedures, complications, clinical trajectory).

- Discharge disposition (location, key services arranged) and the most clinically important medication changes.

- Critical follow-up plan details (scheduled vs recommended appointments, pending results) only if relevant to the readmission story.

4) readmission_leadup_summary

Aim for 2-4 sentences. Focus on the causal chain from discharge to return:

- What went wrong in the gap period (symptom progression, missed follow-ups, med access issues, support gaps---only what is documented).

- How the patient returned (ED vs direct admit, who directed) and the acute trigger (key symptoms/labs/vitals).

5) why_readmission_may_have_occurred

Provide a reasoned analysis that explicitly considers these lenses:

- Discharge planning / care coordination / follow-up (e.g., discharge setting fit, follow-up timing/scheduling, handoff quality, pending results).

- Diagnostic / medication / monitoring (e.g., missed diagnosis, inadequate treatment, medication safety/access, insufficient monitoring or symptom management).

- Patient education / social determinants / goals of care (e.g., understanding of plan, ability to self-manage, support services, ACP issues).

- Appropriate utilization vs unavoidable (e.g., could this have been obs/outpatient vs truly unavoidable due to disease severity).

For each plausible contributor:

- Cite the strongest supporting evidence from the notes and other data (e.g., "Discharge summary note: ...", "Readmission H&P note: ...") in short phrases.

- State 1-3 concrete prevention opportunities that follow logically from the evidence, but only if supported.

6) gaps_identified

Were there actionable care gaps or process failures during the index hospitalization, transition, or gap period that could contribute to readmission? "true" or "false". Note: identifying gaps does NOT necessarily mean the readmission was preventable --- that is judged separately below.

7) gaps_identified_explanation

If gaps were identified, describe each specific gap and the evidence for it. Otherwise leave blank.

8) preventable

A readmission is preventable only if BOTH: (a) a care gap or system failure existed, AND (b) correcting it would have reasonably prevented or delayed the readmission given the patient's clinical trajectory, engagement, and social circumstances. The gap must be plausibly causal, not merely present.

When a patient AMA'd, declined recommended services, or was not engaging with the care team, this substantially reduces preventability even if the system could have done more. The question is whether the patient would have accepted and benefited from improved care.

Options: "true", "false", or "uncertain".

Examples:

- "true": CHF patient readmitted for volume overload; diuretics not uptitrated at discharge despite elevated BNP, no 7-day follow-up scheduled. Patient was engaged and seemed to be adherent with prescribed meds after discharge. System gap with plausible causal link.

- "false": Patient with active substance use AMA'd, declined addiction resources, readmitted with recurrent symptoms driven by ongoing use. Care gaps exist but patient non-engagement makes readmission unavoidable regardless.

- "uncertain": Patient readmitted with surgical site infection; follow-up was at 2 weeks instead of 1. Earlier detection was possible but infection may have been seeded before discharge.

9) preventable_explanation

2-4 sentences explaining your preventability judgment. If gaps were identified but you judged the readmission not preventable, explain why the gaps were not plausibly causal.

10) readmission_primary_category

Which category does the primary reason for readmission best fit into? Choose exactly one of:

- "discharge_planning" --- Discharge planning/instructions, care team coordination, follow-up appointments

- "diagnostic_medication_monitoring" --- Diagnostic, medication, or monitoring causes

- "patient_education_sdoh" --- Patient education, social determinants of health, advance care planning

- "unavoidable" --- Readmission was unavoidable

- "other" --- Other category not listed

11) readmission_primary_category_other

If you selected "other", provide a brief description of the category. Otherwise set this to null.

\# PATIENT DATA

\## Basics

Here is a JSON object with the patient and admission identifiers:

{{ patient_info_json }}

Use this SQLite DB filename: {{ sqlite_filename }}

This is a multi-patient database. Every query must filter by person_id.

Database schema:

- visits: person_id, visit_occurrence_id, visit_start_datetime, visit_end_datetime, visit_concept_name, care_site_name, provider_name, provider_specialty_concept_name

- procedures: person_id, procedure_occurrence_id, visit_occurrence_id, procedure_datetime, procedure_concept_name, procedure_source_value, provider_name, provider_specialty_concept_name

- results: person_id, measurement_id, visit_occurrence_id, measurement_datetime, measurement_name, result_value

- notes: person_id, note_id, visit_occurrence_id, note_datetime, note_title, provider_name, provider_specialty_concept_name, note_text

\## Notes

Some important note_title values: "consults", "progress notes", "imaging", "pathology".

Notes can be very long. Start by sampling only the beginning and end of each note.

Keep early triage queries small (for example LIMIT 20-50), then expand only if needed.

Useful pattern:

`SELECT note_id, note_datetime, note_title,

SUBSTR(note_text, 1, 800) AS note_start,

CASE

WHEN LENGTH(note_text) <= 800 THEN NULL

WHEN LENGTH(note_text) <= 1600 THEN SUBSTR(note_text, 801)

ELSE SUBSTR(note_text, LENGTH(note_text) - 799, 800)

END AS note_end

FROM notes

WHERE person_id = <person_id>

AND note_datetime BETWEEN <start_datetime> AND <end_datetime>

ORDER BY note_datetime;`

Then fetch full note_text only for high-value notes.

\## Results

Culture results are in the results and notes tables. The results table has structured organism identification in the measurement_name and result_value columns. The notes table contains free-text antibiotic sensitivity results in the note_text column, under the note_title 'microbiology culture'.

\# MANDATORY FIRST STEP

Before any other analysis queries, run these two note retrieval queries:

1) First-visit H&P, discharge summary, and discharge instructions

`SELECT note_id, note_datetime, note_title, provider_name, note_text

FROM notes

WHERE person_id = <person_id>

AND visit_occurrence_id = <first_visit_occurrence_id>

AND note_title IN ('h&p', 'discharge summary', 'discharge instructions', 'discharge instr - other info')

ORDER BY note_datetime;`

2) Second-visit ED provider notes, H&P, discharge summary

`SELECT note_id, note_datetime, note_title, provider_name, note_text

FROM notes

WHERE person_id = <person_id>

AND visit_occurrence_id = <second_visit_occurrence_id>

AND note_title IN ('ed provider notes', 'h&p', 'discharge summary')

ORDER BY note_datetime;`

If one of these queries returns no rows, report that in `data_quality_explanation` and continue with other available evidence.

\# DATA EXPLORATION STRATEGY

After the mandatory first step, do a deep dive into the first admission and time between the admissions, focusing on items related to the reason for readmission and potential care gaps that contributed. For instance, if the patient was readmitted with a worsening wound:

- Read consult notes from the first admission related to wound care to make sure the consultants' recommendations were followed

- Read any radiology reports that address the area of the wound (note_title "imaging")

- Read microbiology reports of any wound cultures

- Construct a timeline of the gap between admissions. For this case, read post-discharge home health notes related to the wound, and determine whether the patient took discharge antibiotics as prescribed.

- Look at other items as indicated, like procedures, lab results, and events in the second admission

\# BACKGROUND ON STANFORD QUALITY INITIATIVES

\## Access to outpatient care

Common improvement opportunities seen in readmission provider surveys have been access to outpatient specialist medical care and shorter time to follow-up.

For high readmission risk patients, PCP visit within 7 days recommended. For others, within 14 days. Schedule appointments before discharge whenever possible. Schedule specialty follow-up too (diabetes education, hepatology, oral anticoagulation clinic, oncology, etc.). Discharge summary should list a specific reason for follow-up ("follow up on diuretic dose and daily weights").

Before Dec. 2025, inpatient team had to follow a different process to get patient a visit with internal vs external PCP. After that date, there is a single referral to primary care order in Epic that streamlines the process. The clinical advice service calls patients after discharge to ensure they have an upcoming appointment with their PCP. Getting appointments for patients whose outpatient insurance does not cover Stanford is a continuing gap.

For oncology patients, the Cancer Transitional Care Clinic offers video visits for patients recently discharged from an unplanned admission. Currently deployed for BMT, hematology, oncology, GYN oncology, and GI surgical oncology. Exclusions are hospice and patients not yet established in the Stanford Cancer Center in Palo Alto.

Patients should be provided with the Clinical Advice Service phone number, which is available 24/7 and is staffed by nurses.

\## Medications

Doctors instructed to start discharge planning within 24 hours of admission. Pharmacist helps ensure outpatient meds will be available (Medication Access Program). Transitions of Care pharmacist counseling on discharge is available for patients with many medication changes or high risk of readmission.

\## Other

For patients who were already readmitted, a BPA banner appears in Epic. This should trigger the primary care team to involve multiple teams: case management to coordinate the post-acute plan; social work to connect to community supports; pharmacy to address medication-related issues; nursing to ensure discharge education using teach-back method.

Inpatient team should set goals of care expectations with patient and do advance care planning.

For unhoused patients who live in Santa Clara County or San Mateo County, they should be provided with the phone number of county programs that help with homeless healthcare.

Patients with a preferred language other than English should ideally receive discharge instructions in their preferred language.

\# IMPORTANT RULES

- This is a single-turn conversation. Do not ask clarifying questions.

- If a contributing factor is plausible but not directly supported by evidence, label it as "possible but unconfirmed".

- Output only the final JSON, wrapped in ```json ... ```.

#### AI chart-review prompt for exploratory expanded-cohort analysis

You are a physician-quality reviewer analyzing patients who had readmission to a hospital within 30 days of discharge. Your job: extract the key clinical story, the transition-of-care details, and plausible contributors to the readmission. This will be used later to help modify processes, improve adherence to existing processes, and reduce readmissions. You will analyze one patient at a time.

\# WHAT TO INCLUDE

You will output JSON with exactly eighteen top-level fields:

1) "both_true_inpatient_admissions" (string: "true" or "false")

2) "both_true_inpatient_admissions_explanation" (string)

3) "data_quality" (string or null: "high", "medium", or "low"; null if both_true_inpatient_admissions is "false")

4) "data_quality_explanation" (string or null)

5) "index_hospitalization_summary" (string or null)

6) "readmission_leadup_summary" (string or null)

7) "care_gap_identified" (boolean or null: true or false)

8) "care_gap_primary_category" (string or null: one of the care-gap category options, null if no care gaps, or null if both_true_inpatient_admissions is "false")

9) "care_gap_primary_category_other" (string or null: only populated if primary category is "other")

10) "care_gap_other_categories" (array of strings or null: zero or more additional care-gap category options)

11) "care_gap_other_categories_other" (string or null: only populated if other categories include "other")

12) "care_gap_discussion" (string or null)

13) "preventable_rating" (integer or null: 1 to 5)

14) "preventable_confidence" (string or null: "high", "medium", or "low")

15) "preventable_explanation" (string or null)

16) "actionable_improvement_opportunity" (string or null: "no", "individual_case_feedback", "local_workflow_change", or "system_change")

17) "improvement_opportunity_owners" (array of strings or null: zero or more owner options)

18) "suggested_intervention" (string or null)

1) both_true_inpatient_admissions

Determine whether BOTH the index hospitalization and the readmission qualify as true inpatient admissions AND the second admission was unplanned. To qualify, each encounter must be a genuine inpatient stay --- not an observation stay, an outpatient encounter (e.g., hospital-based radiology, infusion, or clinic visit), an ED visit that did not convert to inpatient admission, or a same-day procedure without overnight stay. The second admission must not be a planned admission for scheduled treatment, placement, direct transfer to another hospital/service, or other prearranged continuation of care. Use admission/discharge notes and length of stay to make this determination. Options: "true" or "false". If "false", then do not continue with the analysis and mark fields 3-18 as null.

2) both_true_inpatient_admissions_explanation

1-2 sentences explaining your determination.

3) data_quality

Rate the completeness of the input data available for your analysis. This reflects how much source material you had to work with, not your confidence in your conclusions.

- "high": Index discharge summary and readmission H&P/ED provider note are both present and substantive; SQLite database contains meaningful gap-period notes, labs, or other relevant data.

- "medium": One key document (discharge summary or readmission H&P/ED provider note) is missing, blank, or a brief stub; OR the SQLite database has limited gap-period data.

- "low": Multiple key inputs are missing or blank (e.g., both discharge summary and H&P/ED provider notes are absent/stubs, or the SQLite database has minimal useful data).

4) data_quality_explanation

1-2 sentences listing specifically what was missing or degraded. If data quality is high, briefly state what was available.

5) index_hospitalization_summary

Aim for 3-5 sentences. Cover:

- Primary diagnosis, key comorbidities affecting discharge risk.

- Hospital course highlights (major procedures, complications, clinical trajectory).

- Discharge disposition (location, key services arranged) and the most clinically important medication changes.

- Critical follow-up plan details (scheduled vs recommended appointments, pending results) only if relevant to the readmission story.

6) readmission_leadup_summary

Aim for 2-4 sentences. Focus on the causal chain from discharge to return:

- What went wrong in the gap period (symptom progression, missed follow-ups, med access issues, support gaps---only what is documented).

- How the patient returned (ED vs direct admit, who directed) and the acute trigger (key symptoms/labs/vitals).

7) care_gap_identified

Were there care gaps or process failures during the index hospitalization, transition, or gap period that would be of interest to the hospital medicine quality team? Boolean: true or false.

8) care_gap_primary_category

Which category do the care gaps best fit into? Choose exactly one of:

- "discharge_planning" --- Discharge planning/instructions, care team coordination, follow-up appointments

- "diagnostic_medication_monitoring" --- Diagnostic, medication, or monitoring causes

- "patient_education_sdoh" --- Patient education, social determinants of health, advance care planning

- "other" --- Other category not listed

- null - No care gaps identified

9) care_gap_primary_category_other

If you selected "other", provide a brief description of the category. Otherwise set this to null.

10) care_gap_other_categories

List any additional care-gap categories beyond `care_gap_primary_category`. Use an array containing zero or more of the same category strings: "discharge_planning", "diagnostic_medication_monitoring", "patient_education_sdoh", "other". Do not repeat the primary category. If there are no care gaps or no secondary categories, use an empty array.

11) care_gap_other_categories_other

If care_gap_other_categories contains "other", provide a brief explanation. Otherwise set this to null.

12) care_gap_discussion

Provide a reasoned analysis that explicitly considers these lenses:

- Discharge planning / care coordination / follow-up (e.g., discharge setting fit, follow-up timing/scheduling, handoff quality, pending results).

- Diagnostic / medication / monitoring (e.g., missed diagnosis, inadequate treatment, medication safety/access, insufficient monitoring or symptom management).

- Patient education / social determinants / goals of care (e.g., understanding of plan, ability to self-manage, support services, ACP issues).

- Appropriate utilization vs unavoidable (e.g., could this have been obs/outpatient vs truly unavoidable due to disease severity).

13) preventable_rating

Rate the preventability of this readmission on a 5-point Likert scale. A readmission is preventable if (a) a care gap or system failure existed, AND (b) correcting it would have reasonably prevented or delayed the readmission.

1: Definitely Not Preventable. No care gaps identified; readmission was an unavoidable progression of disease or a new acute event.

2: Probably Not Preventable. Minor gaps found, but unlikely to have caused the readmission given the clinical trajectory.

3: Possibly Preventable. Actionable care gaps identified with a plausible but uncertain causal link; success not guaranteed even with perfect care.

4: Probably Preventable. Clear care gaps identified that likely directly contributed to the readmission.

5: Definitely Preventable. Glaring process failure or error; highly avoidable with standard-of-care transition/management.

When a patient left AMA, declined recommended services, or was not engaging with the care team, assess whether that barrier substantially contributed to the readmission. If it did, the readmission should generally not be considered preventable and the preventable_rating should not be greater than 2, even if the system could have done more. Do not apply this cap if the patient-initiated barrier appears to have been driven primarily by a remediable system issue, such as inadequate communication, untreated symptoms, unsafe discharge planning, or failure to address access/social needs.

Examples:

- preventable_rating=5, preventable_confidence="high": CHF patient readmitted for volume overload; diuretics not uptitrated at discharge despite elevated BNP, no 7-day follow-up scheduled. Patient was engaged and adherent.

- preventable_rating=1, preventable_confidence="high": Patient with active substance use AMA'd, declined resources, readmitted with recurrent symptoms driven by ongoing use.

- preventable_rating=3, preventable_confidence="low": Patient readmitted with surgical site infection; follow-up was at 2 weeks instead of 1. Earlier detection is plausible, but infection may have been seeded before discharge and might have required readmission regardless.

14) preventable_confidence

How confident are you in your specific 1-5 rating?

- "high": Clear evidence and causal chain; very little doubt about the appropriate rating.

- "medium": Evidence leans one way, but missing details or clinical complexity make the specific rating a judgment call.

- "low": Weak/ambiguous evidence; the rating is a forced best-guess and could easily shift with more data.

15) preventable_explanation

2-4 sentences explaining your preventability rating. If gaps were identified but the rating is low, explain why the gaps were not plausibly causal. If confidence is "low", explicitly name the 1-2 missing or ambiguous facts driving uncertainty.

16) actionable_improvement_opportunity

Options: "no", "individual_case_feedback", "local_workflow_change", "system_change"

- "no": No specific actionable change is supported.

- "individual_case_feedback": Case-specific feedback to one clinician/team; no durable workflow change is implied.

- "local_workflow_change": A change to a service/team workflow, checklist, handoff, discharge process, pharmacy review, scheduling practice, or monitoring process could reduce similar events.

- "system_change": A cross-team, institution-level, IT, policy, staffing, resource, or access issue needs broader operational change.

17) improvement_opportunity_owners

Array of strings. Options: "hospitalist_service", "case_management_social_work", "pharmacy", "nursing", "ED", "outpatient_clinic", "IT", or another team name. If none, use an empty array.

18) suggested_intervention

Brief description of the suggested intervention, or null.

\# CROSS-FIELD CONSISTENCY RULES

- If `both_true_inpatient_admissions` is "false", explain why and set fields 3-18 to null.

- Preventability ratings 3-5 require at least one documented care gap with a plausible causal link to the readmission. Ratings 4-5 require a clear causal link.

- Rating 2 may include minor gaps, but the explanation must state why they were unlikely to cause the readmission.

- Do not assign an improvement owner or intervention unless `actionable_improvement_opportunity` is not "no".

\# PATIENT DATA

\## Basics

Here is a JSON object with the patient and admission identifiers:

{{ patient_info_json }}

Use this SQLite DB filename: {{ sqlite_filename }}

This is a multi-patient database. Every query must filter by person_id.

Database schema:

- visits: person_id, visit_occurrence_id, visit_start_datetime, visit_end_datetime, visit_concept_name, care_site_name, provider_name, provider_specialty_concept_name

- procedures: person_id, procedure_occurrence_id, visit_occurrence_id, procedure_datetime, procedure_concept_name, procedure_source_value, provider_name, provider_specialty_concept_name

- results: person_id, measurement_id, visit_occurrence_id, measurement_datetime, measurement_name, result_value

- notes: person_id, note_id, visit_occurrence_id, note_datetime, note_title, provider_name, provider_specialty_concept_name, note_text

\## Notes

Some important note_title values: "consults", "progress notes", "imaging", "pathology".

STARR-OMOP date handling: structured data dates are jittered by the same amount for each patient, so structured visit/procedure/result dates can be compared internally for that patient. Dates mentioned inside note_text are usually jittered by the same amount as the structured dates, but the date recognition is imperfect. So if a date in a note appears offset from structured data or other notes, do not treat that mismatch by itself as evidence that the note is erroneous.

Notes can be very long. Start by sampling only the beginning and end of each note.

Keep early triage queries small (for example LIMIT 20-50), then expand only if needed.

Useful pattern:

`SELECT note_id, note_datetime, note_title,

SUBSTR(note_text, 1, 800) AS note_start,

CASE

WHEN LENGTH(note_text) <= 800 THEN NULL

WHEN LENGTH(note_text) <= 1600 THEN SUBSTR(note_text, 801)

ELSE SUBSTR(note_text, LENGTH(note_text) - 799, 800)

END AS note_end

FROM notes

WHERE person_id = <person_id>

AND note_datetime BETWEEN <start_datetime> AND <end_datetime>

ORDER BY note_datetime;`

Then fetch full note_text only for high-value notes.

\## Results

Culture results are in the results and notes tables. The results table has structured organism identification in the measurement_name and result_value columns. The notes table contains free-text antibiotic sensitivity results in the note_text column, under the note_title 'microbiology culture'.

\# MANDATORY FIRST STEP

Before any other analysis queries, run these three queries:

0) Visit type verification for both encounters

`SELECT visit_occurrence_id, visit_concept_name, visit_start_datetime, visit_end_datetime, care_site_name

FROM visits

WHERE person_id = <person_id>

AND visit_occurrence_id IN (<first_visit_occurrence_id>, <second_visit_occurrence_id>)

ORDER BY visit_start_datetime;`

Use this to help populate the `both_true_inpatient_admissions` fields.

1) First-visit H&P, discharge summary, and discharge instructions

`SELECT note_id, note_datetime, note_title, provider_name, note_text

FROM notes

WHERE person_id = <person_id>

AND visit_occurrence_id = <first_visit_occurrence_id>

AND note_title IN ('h&p', 'discharge summary', 'discharge instructions', 'discharge instr - other info')

ORDER BY note_datetime;`

2) Second-visit ED provider notes, H&P, discharge summary

`SELECT note_id, note_datetime, note_title, provider_name, note_text

FROM notes

WHERE person_id = <person_id>

AND visit_occurrence_id = <second_visit_occurrence_id>

AND note_title IN ('ed provider notes', 'h&p', 'discharge summary')

ORDER BY note_datetime;`

If one of these queries returns no rows, report that in `data_quality_explanation` and continue with other available evidence.

\# DATA EXPLORATION STRATEGY

After the mandatory first step, do a deep dive into the first admission and time between the admissions, focusing on items related to the reason for readmission and potential care gaps that contributed. For instance, if the patient was readmitted with a worsening wound:

- Read consult notes from the first admission related to wound care to make sure the consultants' recommendations were followed

- Read any radiology reports that address the area of the wound (note_title "imaging")

- Read microbiology reports of any wound cultures

- Construct a timeline of the gap between admissions. For this case, read post-discharge home health notes related to the wound, and determine whether the patient took discharge antibiotics as prescribed.

- Look at other items as indicated, like procedures, lab results, and events in the second admission

\# BACKGROUND ON STANFORD QUALITY INITIATIVES

\## Access to outpatient care

Common improvement opportunities seen in readmission provider surveys have been access to outpatient specialist medical care and shorter time to follow-up.

For high readmission risk patients, PCP visit within 7 days recommended. For others, within 14 days. Schedule appointments before discharge whenever possible. Schedule specialty follow-up too (diabetes education, hepatology, oral anticoagulation clinic, oncology, etc.). Discharge summary should list a specific reason for follow-up ("follow up on diuretic dose and daily weights").

Before Dec. 2025, inpatient team had to follow a different process to get patient a visit with internal vs external PCP. After that date, there is a single referral to primary care order in Epic that streamlines the process. The clinical advice service calls patients after discharge to ensure they have an upcoming appointment with their PCP. Getting appointments for patients whose outpatient insurance does not cover Stanford is a continuing gap.

For oncology patients, the Cancer Transitional Care Clinic offers video visits for patients recently discharged from an unplanned admission. Currently deployed for BMT, hematology, oncology, GYN oncology, and GI surgical oncology. Exclusions are hospice and patients not yet established in the Stanford Cancer Center in Palo Alto.

Patients should be provided with the Clinical Advice Service phone number, which is available 24/7 and is staffed by nurses.

\## Medications

Doctors instructed to start discharge planning within 24 hours of admission. Pharmacist helps ensure outpatient meds will be available (Medication Access Program). Transitions of Care pharmacist counseling on discharge is available for patients with many medication changes or high risk of readmission.

\## Other

For patients who were already readmitted, a BPA banner appears in Epic. This should trigger the primary care team to involve multiple teams: case management to coordinate the post-acute plan; social work to connect to community supports; pharmacy to address medication-related issues; nursing to ensure discharge education using teach-back method.

Inpatient team should set goals of care expectations with patient and do advance care planning.

For unhoused patients who live in Santa Clara County or San Mateo County, they should be provided with the phone number of county programs that help with homeless healthcare.

Patients with a preferred language other than English should ideally receive discharge instructions in their preferred language.

\# IMPORTANT RULES

- This is a single-turn conversation. Do not ask clarifying questions.

- Citations are mandatory for any mention of clinical facts from the EMR record in any free-text field.

- Use the following structured reference formats:

- `[note:ID]` for citations from the notes table (note_id).

- `[result:ID]` for citations from the results table (measurement_id).

- `[proc:ID]` for citations from the procedures table (procedure_occurrence_id).

- `[visit:ID]` for citations from the visits table (visit_occurrence_id).

- Place these references immediately after the relevant claim or at the end of the sentence.

- If a contributing factor is plausible but not directly supported by evidence, label it as "possible but unconfirmed".

- Output only the final JSON, wrapped in ```json ... ```.

#### Chart review scoring rubric

| Item | Instructions |
| --- | --- |
| Overall quality | 1 = Poor: confusing, incomplete, or low-value review  3 = Adequate: generally reasonable, with some omissions or weak points  5 = Excellent: clear, clinically sensible, well-supported, and high-value |
| Factuality | 1 = Major unsupported or incorrect claims  3 = Mostly accurate, but some statements are weakly supported, overstated, or slightly questionable  5 = Accurate, well-supported, and appropriately cautious throughout |
| Actionability | 1 = Not useful for quality improvement; little clear takeaway  3 = Some useful ideas, but limited specificity or relevance  5 = Clearly useful; identifies specific, relevant opportunities or conclusions that could inform action |
| Guessed chart reviewer identity | Human / AI |
| Comments | Optional |

#### Free-text similarity scoring scale

| Score | Label | Description |
| --- | --- | --- |
| 1 | Fundamentally different | The entries have little meaningful overlap or they conflict on the main story, conclusion, or causal reasoning. |
| 2 | Weakly similar | The entries share only a broad topic or a few surface details, but important clinical content, reasoning, or conclusions differ substantially. |
| 3 | Moderately similar | The entries overlap on the main theme, but there are noticeable omissions, additions, or emphasis differences that change some important aspects of the field. |
| 4 | Strongly similar | The entries convey the same main clinical story or reasoning, with only minor differences in wording, specificity, or secondary details. |
| 5 | Near-equivalent | The entries are essentially paraphrases at the meaning level, with no meaningful clinical or interpretive differences. |

### Supplemental Results

#### Chart review patterns

**Supplemental Table S1.** Patterns of structured field answers. Physician #1 reviewed all 20 charts; physicians #2 and #3 reviewed 10 charts each.

| Item | AI | Physician #1 | Physician #2 | Physician #3 |
| --- | --- | --- | --- | --- |
| Readmission preventable | 9/20 | 9/20 | 7/10 | 3/10 |
| Readmission primary category |  |  |  |  |
| Diagnostic/ medication/ monitoring | 8/20 | 2/20 | 1/10 | 2/10 |
| Discharge planning | 3/20 | 3/20 | 4/10 | 1/10 |
| Patient education/ social determinants of health | 1/20 | 0/20 | 2/10 | 2/10 |
| Unavoidable | 8/20 | 12/20 | 3/10 | 4/10 |
| Other | 0/20 | 3/20 | 0/10 | 1/10 |

**Supplemental Table S2.** Mean number of words in AI versus human free-text answers.

| **Field** | **AI mean no. words (IQR)** | **Human mean no. words (IQR)** | **p value*** |
| --- | --- | --- | --- |
| Index hospitalization summary | 90.7 (85.0, 96.2) | 66.3 (53.2, 76.1) | <0.001 |
| Readmission lead-up summary | 69.7 (64.8, 74.5) | 45.5 (39.1, 47.1) | <0.001 |
| Why readmission may have occurred | 165.7 (147.0, 173.25) | 37.7 (23.5, 41.0) | <0.001 |
| Preventability explanation | 71.2 (64.8, 78.3) | 44.8 (33.8, 52.1) | <0.001 |
| Overall | 99.3 (93.3, 103.0) | 48.6 (39.1, 55.7) | <0.001 |

* Two-sided exact paired sign-flip permutation test on the patient-level, AI vs. mean of two humans

**Supplemental Table S3.** Results for selected patients. Some details have been changed to preserve anonymity.

| Patient summary | AI review summary | Physician review summary | Evaluator overall ratings (1-5) | Evaluator comments |
| --- | --- | --- | --- | --- |
| Elderly woman with cerebral amyloid angiopathy (CAA), atrial fibrillation, CAD, severe dysphagia, and chronic SIADH was admitted for fever, hypovolemia, and symptomatic hyponatremia (Na 125). Fevers attributed to hypovolemia and possible aspiration pneumonitis. Home sodium chloride dose increased and urea packets started. Discharged home on a pureed diet. Five days after discharge, the patient developed recurrent fever and an episode of unresponsiveness and was readmitted. Found to have recurrent hyponatremia (Na 127) with positive blood culture, which was later found to be from a contaminant. | Classified as preventable. Follow-up PCP appointment not scheduled at time of discharge. And "the patient was prescribed large 1g salt tablets and non-crushable extended-release medications (e.g., Metoprolol succinate) despite documented severe dysphagia and a pureed diet. Difficulty administering these likely caused poor adherence, worsening hyponatremia, and lethargy that prompted the OSH ED visit." Should use a transition of care pharmacist in the future for patients with severe dysphagia to ensure liquid/crushable meds. | One physician reviewer felt the readmission was unavoidable due to the positive blood culture. The other felt it was preventable since the patient was clinically stable, had end-stage dementia with aspiration events expected, and a full inpatient admission was overutilization. Goals of care and family member education during the first admission could have helped prevent the readmission. | AI: 4  Physicians: 2, 5 | Evaluator was impressed by the AI noticing the issue with the patient’s dysphagia and specific medications, but did not think this was the main driver of readmission. All three reviewers made valid points. This case highlights the complex causes of readmissions and difficulty assigning a single cause or preventability rating. |
| Middle-aged woman with complex urologic history admitted for pyelonephritis. She was restarted on chronic buprenorphine/naloxone. Hypothyroid with TSH >200 mIU/L, kept on home dose of levothyroxine, had likely not been taking this. Discharged without PCP appointment (was later made for 26 days post-discharge) or bowel regimen. Readmitted a week after discharge with nausea, vomiting, constipation, PO intolerance, acute kidney injury. | Classified as preventable. Readmission driven by constipation from resumed buprenorphine use and inadequately treated hypothyroidism. Patient did not receive recommended 7-day follow-up. Should have had bowel prophylaxis. Should have had levothyroxine increase or endocrinology consult. | Physicians agreed readmission was preventable due to the patient being discharged without counseling on opioid-induced constipation and without any prophylactic laxatives or bowel regimen. Did not mention TSH. | AI: 4  Physicians: 3, 4 | The evaluator agreed with the AI system that the very high TSH was relevant. But since the patient likely been noncompliant with levothyroxine and was not in myxedema coma, the reviewer disagreed with the AI system and felt that it was reasonable to continue the home dose of levothyroxine and not consult endocrinology. |
| Elderly man with advanced dementia, heart failure, pulmonary hypertension admitted with sacral pressure ulcers and concern for neglect. Discharged to child’s care with home health. Readmitted with hypercarbic respiratory failure and aspiration pneumonia, transitioned to comfort care. | Classified as unpreventable due to terminal disease progression. Flagged two care gaps:  1) Outpatient follow-up on day 18, outside the recommended 7-day high risk window  2) The post-discharge phone call failed because the hospital had an incorrect contact number on file | One physician classified as unpreventable due to patient’s overall trajectory, and readmission causes being unrelated to first admission acute issues. One classified as preventable because early outpatient visit could have triggered home hospice. | AI: 5  Physicians: 4, 4 | The evaluator was impressed with the AI noticing the detail about the incorrect phone number on file, even though it was not a main driver of the readmission. |

#### Similarity between reviews

*Structured fields*

For key structured fields (whether the readmission was preventable, and readmission primary category), agreement was generally low between all case reviewers (kappa statistic of -0.10 to 0.25, Supplemental Table S4). Human-human agreement for whether a readmission was preventable was 45% (9/20), compared to 62.5% (25/40) for human-AI agreement.

*Free-text fields*

We had an LLM rate how similar each pair of free-text responses was for each patient on a 5-point Likert scale. To validate this LLM-as-a-judge approach, we had a physician reviewer also do this scoring for a subset of responses. The LLM and physician scores showed good agreement (n=120 text pairs, quadratic weighted kappa 0.61, within-1 agreement 89%), so the LLM was used for subsequent ratings.

Based on the LLM ratings, mean similarity score for AI-human text pairs was 2.93, compared to 2.69 for human-human pairs. AI-human similarity scores were statistically higher than human-human similarity scores for two of four key free-text fields, index hospitalization summary and readmission lead-up summary (Supplemental Table S5).

**Supplemental Table S4.** Similarity scores for key structured fields.

| **Field** | **AI-human agreement (%)** | **AI-human agreement Kappa** | **Human-human agreement (%)** | **Human-human agreement Kappa** | **p value for difference between AI-human vs. human-human similarity*** |
| --- | --- | --- | --- | --- | --- |
| Readmission preventable | 25/40 (62.5%) | 0.25 | 9/20 (45%) | -0.10 | 0.17 |
| Readmission primary category | 16/40 (40%) | 0.17 | 6/20 (30%) | 0.04 | 0.53 |

* Two-sided exact paired sign-flip permutation test on patient-level differences

**Supplemental Table S5.** Free-text field similarity scores.

| **Field** | **Mean AI-human similarity score** | **Mean human-human similarity score** | **p value*** |
| --- | --- | --- | --- |
| Index hospitalization summary | 3.60 | 3.30 | 0.02 |
| Readmission lead-up summary | 3.23 | 2.75 | 0.015 |
| Why readmission may have occurred | 2.33 | 2.55 | 0.36 |
| Preventable explanation | 2.58 | 2.15 | 0.135 |

* Two-sided exact paired sign-flip permutation test on patient-level differences

#### Exploratory expanded-cohort analysis

**Supplemental Table S6.** Patient characteristics for the exploratory expanded-cohort analysis. CCSR categories and days from discharge to readmission are for all 100 readmission events; other items are for the first admission per patient (81 admissions).

| Variable | Median (Q1, Q3) or count (percentage) |
| --- | --- |
| Age | 63.0 (44.0, 74.0) |
| **Sex** |  |
| Female | 38 (47%) |
| Male | 43 (53%) |
| **Race/ethnicity** |  |
| Hispanic | 28 (35%) |
| Non-Hispanic White | 26 (32%) |
| Non-Hispanic Black | 12 (15%) |
| Non-Hispanic Asian | 8 (10%) |
| Unknown | 7 (9%) |
| **Insurance status** |  |
| Medicare | 22 (27%) |
| Medicaid | 44 (54%) |
| Commercial | 14 (17%) |
| Unknown | 1 (1%) |
| **Index admission CCSR* diagnosis category** |  |
| Blood | 1 (1%) |
| Circulatory | 13 (13%) |
| Digestive | 16 (16%) |
| Endocrine, nutritional, metabolic | 13 (13%) |
| External causes of morbidity | 1 (1%) |
| Genitourinary | 9 (9%) |
| Infectious | 3 (3%) |
| Injury/poisoning | 6 (6%) |
| Mental, behavioral, neurodevelopmental | 3 (3%) |
| Musculoskeletal | 2 (2%) |
| Neoplasms | 2 (2%) |
| Respiratory | 19 (19%) |
| Skin | 4 (4%) |
| Symptoms, signs, abnormal findings | 8 (8%) |
| **Readmission CCSR diagnosis category** |  |
| Blood | 1 (1%) |
| Circulatory | 9 (9%) |
| Digestive | 16 (16%) |
| Endocrine, nutritional, metabolic | 14 (14%) |
| Genitourinary | 10 (10%) |
| Infectious | 4 (4%) |
| Injury/poisoning | 7 (7%) |
| Mental, behavioral, neurodevelopmental | 8 (8%) |
| Musculoskeletal | 2 (2%) |
| Nervous system | 2 (2%) |
| Respiratory | 17 (17%) |
| Skin | 2 (2%) |
| Symptoms, signs, abnormal findings | 8 (8%) |
| Days from discharge to readmission | 8 (5, 13) |

*Agency for Healthcare Research & Quality Clinical Classifications Software Refined

**Supplemental Table S7.** Primary care gap categories for the exploratory expanded-cohort analysis.

| **Gap** | **Number** |
| --- | --- |
| Discharge planning | 51 |
| Diagnostic/medication/monitoring | 36 |
| Patient education/social determinants of health | 6 |
| Other | 1 |
| None | 6 |

**Supplemental Table S8.** Actionability categories for improvement opportunities in the exploratory expanded-cohort analysis.

| **Category** | **Number** |
| --- | --- |
| Local workflow change | 74 |
| System change (institution-level) | 11 |
| Individual case feedback | 8 |
| No actionable opportunity | 7 |

**Supplemental Table S9.** Stakeholder ownership distribution for improvement opportunities in the exploratory expanded-cohort analysis (one case could have more than one stakeholder).

| **Stakeholder** | **Number** |
| --- | --- |
| Hospitalist service | 84 |
| Pharmacy | 44 |
| Case management/social work | 41 |
| Nursing | 14 |
| Outpatient clinic | 13 |
| Total | 196 |

**Supplemental Table S10.** Most common codes from qualitative analysis of the AI chart summaries in the exploratory expanded-cohort analysis (codes assigned to at least 5 cases are shown).

| Code | Number of cases |
| --- | --- |
| **Category: Readmission drivers** |  |
| Discharge planning: follow-up not scheduled | 35 |
| Diagnostic/therapeutic problems: inadequate treatment | 32 |
| Medication safety: discharge-order errors | 20 |
| Medication safety: inadequate monitoring | 12 |
| Diagnostic/therapeutic problems: prescription access issues | 11 |
| Discharge planning: unclear discharge instructions | 10 |
| Discharge planning: discharged too soon | 6 |
| Discharge planning: inappropriate discharge location | 6 |
| Medication safety: prophylactic medication omitted | 5 |
| Symptom/disease monitoring: lack of disease monitoring | 5 |
| **Category: Guideline adherence gaps** |  |
| Primary care follow-up not scheduled at discharge | 30 |
| Predischarge specialist scheduling failure | 18 |
| No transitions-of-care pharmacist review documented | 11 |
| Medication access program omitted | 7 |
| **Category: Prevention opportunities** |  |
| Predischarge follow-up secured | 29 |
| Medication reconciliation and affordability support | 28 |
| Enhanced patient/caregiver education | 9 |
| Inpatient workup completion | 9 |
| Enhanced outpatient monitoring | 8 |
| Post-acute level-of-care optimization | 8 |
| **Category: Patient factors** |  |
| Behavioral/psychiatric: active substance use | 16 |
| Behavioral/psychiatric: self-directed discharge | 16 |
| Clinical/functional: complex regimen burden | 14 |
| Clinical/functional: frailty or fall risk | 13 |
| Behavioral/psychiatric: intentional nonadherence | 11 |
| Clinical/functional: cognitive impairment | 10 |
| Behavioral/psychiatric: severe psychiatric condition | 9 |
| Clinical/functional: terminal illness progression | 8 |
| Social determinants: financial or insurance barriers | 8 |
| Behavioral/psychiatric: refusal of post-acute services | 6 |
